# Point Sensitivity of the Multi-target Stool DNA Test for Colorectal Cancer Screening in Individuals Ages 45-49 Years

**DOI:** 10.1101/2020.09.01.20177568

**Authors:** Michael Domanico, Sandra Statz, Emily Weiser, Barry M Berger, Paul Limburg

## Abstract

**Background:** Most colorectal cancer (CRC) screening tests have not been rigorously studied in younger age groups.

**Aims:** To estimate sensitivity of the multi-target stool DNA (mt-sDNA) test in patients ages 45-49 years.

**Methods:** We identified archived stool samples (Exact Sciences; Madison, WI) from individuals ages 45-49 years who had completed an index colonoscopy and had confirmed diagnoses of CRC or advanced precancerous lesions (APL; defined as high-grade dysplasia, >25% villous morphology, or ≥1 cm in size [conventional adenoma or serrated lesion]). Data annotation referent to potential CRC risk factors, other than age, was limited. Stool samples were collected at least 7 days after the index colonoscopy, prior to lesion excision or treatment. Stool samples were processed and analyzed per established laboratory protocols for the mt-sDNA assay. Mt-sDNA test sensitivity for CRC, APL, and CRC+APL was estimated from the available sample set. Samples were collected from 2010-2013 (NCT01260168) and 2014-2017 (NCT02503631), with sample testing and analysis in 2019.

**Results:** Stool samples were analyzed from 19 eligible subjects, 13 with CRC and 6 with APL. Estimated mt-sDNA test sensitivity for CRC, APL, and CRC+APL were 92%, 83%, and 89%, respectively.

**Conclusions:** In this small pilot study using existing archived stool samples from subjects ages 45-49 years, mt-sDNA test sensitivity was similar to previously reported estimates for individuals ages ≥50 years. These results support the application of mt-sDNA screening to 76 average-risk patients beginning at age 45 years. Larger studies are needed to confirm and extend these findings.

## Introduction

Colorectal cancer (CRC) is the fourth most-diagnosed cancer and second deadliest cancer in the United States.[1] Although CRC incidence and mortality rates in the ≥55-year-old population are decreasing, CRC incidence and mortality rates are increasing in those <55 years of age.[2] In 2020, 17,930 new CRC cases and 3640 deaths are estimated in individuals <50 years-old.[3] The American Cancer Society (ACS) CRC screening guidelines now recommend initiating CRC screening at age 45 years for individuals at average risk for CRC, whereas previous iterations of the guidelines recommended initiating screening at age 50 for most average-risk individuals.[4] The endorsed CRC screening test options remained unchanged in the older and younger populations. However, most endorsed CRC screening tests have not been rigorously evaluated in individuals <50 years of age.

One ACS-recommended CRC screening option is the multi-target stool DNA (mt-sDNA) test. The mt-sDNA test was approved by the US Food and Drug Administration (FDA) in 2014 to screen individuals ≥50 years at average risk for CRC and was granted an FDA label expansion in 2019 to screen average-risk individuals beginning at age 45 years. In a study of nearly 10,000 participants ≥50 years at average risk for CRC, the mt-sDNA test was 92% sensitive for CRC, 42% sensitive for advanced precancerous lesions (APL), and 87% specific, including participants with nonadvanced adenomas (NAA) or negative findings on colonoscopy.[5]

Although existing data do not suggest age-related changes in sensitivity of the mt-sDNA test,[5] further investigation is needed to more fully define the mt-sDNA test performance for average-risk CRC screening in individuals 45-49 years of age. In this study, we examined the point sensitivity of the multi-target stool DNA (mt-sDNA) test, using archived stool samples from patients ages 45-49 years and with confirmed CRC or APL.

## Methods

### Study Samples

The Exact Sciences (Madison, WI) biorepository was queried to identify archived stool samples previously collected (NCT01260168, NCT02503631), prepared for stool DNA testing, and stored at −80°C from individuals ages 45-49 years who underwent an index colonoscopy with confirmed diagnosis of CRC or APL. APL were defined as neoplasia with high-grade dysplasia or ≥25% villous morphology, or conventional adenomas or serrated lesions ≥1 cm in size. Data annotation referent to potential CRC risk factors, other than age, were not systematically collected from subjects with archived stool samples and were, therefore, limited. Stool samples were collected at least 7 days after the index colonoscopy, prior to lesion excision or treatment. Stool samples were processed and analyzed per established laboratory protocols for the mt-sDNA assay. Additionally, data from previously tested stool samples collected from individuals ages 45-49 years were identified and included in the analyses. Stool samples were collected from 2010 to 2013 (NCT01260168) and 2014 to 2017 (NCT02503631) during studies approved by the Copernicus Group Institutional Review Board. Use of the samples for the current study was covered in the original informed consent. Sample testing and retrospective analyses occurred in 2019.

### Measures

Mt-sDNA test point sensitivity estimates for CRC, APL, and advanced colorectal neoplasia (CRC+APL) were defined as the primary study endpoints.

### Statistical Analysis

The number and percent of positive mt-sDNA test results were summarized for each neoplasia outcome of interest. Given the limited availability of archived stool samples from subjects who met the eligibility criteria, this pilot study was not powered for more robust statistical analyses.

## Results

Previously tested and archived samples were available from a total of 19 eligible subjects, including 13 with CRC and 6 with APL. The average size of index lesions was 3 cm; however, lesion size was missing from 3 samples. Mt-sDNA test sensitivity was 92% for CRC, 83% for APL, and 89% for advanced colorectal neoplasia (CRC+APL) (Table 1).

**Table 1.**
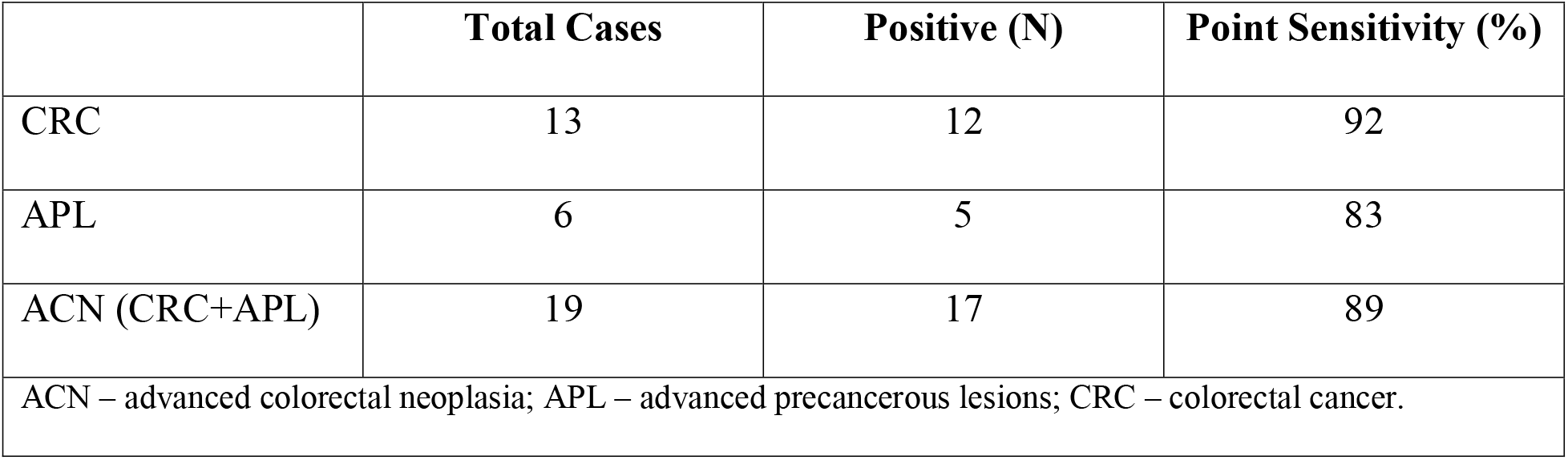
Point Sensitivity of the mt-sDNA Test for Colorectal Cancer, Advanced Precancerous Lesions, and Advanced Colorectal Neoplasia

## Discussion

Data from this relatively small, pilot study using existing, archived stool samples from individuals ages 45-49 years with clinically confirmed CRC or APL demonstrate high point sensitivity of the mt-sDNA test for advanced colorectal neoplasia (89%). While CRC sensitivity at 92% was similar to previously reported estimates for individuals ages 50 years and older, APL sensitivity at 83% exceeded that seen previously in individuals ≥50 years,[5] though this is based on a small, convenience sample of archived stool specimens. This study provides an initial demonstration of mt-sDNA test sensitivity in this younger population, which supports the recent label expansion to extend the application of this test to average-risk CRC screening for individuals ages 45 and older. Of note, a large, prospective clinical trial (NCT03728348) to assess specificity of the mt-sDNA test in individuals 45-49 years-old was recently completed and will further define the mt-sDNA test performance in this younger age group.

### Limitations

The major limitation of this pilot study is the relatively small sample size, which was determined primarily by the availability of existing, archived stool specimen data. Since the mt-sDNA test was originally approved for CRC screening in average-risk patients ≥50 years, stool samples have not been frequently collected from younger patients for molecular marker interrogation in routine clinical practice. As such, more comprehensive data analyses were not feasible for this initial investigation of mt-sDNA test sensitivity due to limited statistical power. Additionally, the average index lesion size in this study (3 cm) was larger than would be expected in a typical CRC screening population. This may overestimate the sensitivity compared to an average-risk screening population. Nonetheless, based on the analyzed samples, mt-sDNA test sensitivity was similar to previously reported data for individuals 50 years and older at average risk for CRC,[5] supporting the expectation of comparable test sensitivity in this younger population, which will be further clarified by forthcoming data from additional studies.

The mt-sDNA test demonstrated high point sensitivity for advanced colorectal neoplasia in stool samples from patients ages 45-49 years, supporting its application as a widely accessible, noninvasive option for average-risk CRC screening in this age group.

## Data Availability

Data are available on reasonable request. Proposals for access to data should be directed to the corresponding author. To gain access, data requestors will need to provide a methodologically sound proposal and sign a data access agreement. Researchers are required to obtain necessary IRB/EC approvals or waivers as applicable to conduct research.

## Other Acknowledgments

Medical writing and editorial support was provided by Rebecca K Swartz, PhD, an employee of Exact Sciences (Madison, WI).

